# Retinal and Optic Nerve Lesions Correspond to Amyloid in Autosomal Dominant Alzheimer’s Disease

**DOI:** 10.1101/2025.01.21.25319904

**Authors:** Amir H Kashani, Maya Koronyo-Hamaoui, Yosef Koronyo, Haoshen Shi, Muhammed Alluwimi, Maxwell Singer, Abhay Sagare, Debra Hawes, Anna Tang, Xuejuan Jiang, Ana Collazo Martinez, Fred N. Ross-Cisneros, Alfredo A. Sadun, John M. Ringman

## Abstract

Autosomal dominant Alzheimer’s disease (ADAD) is a rare form of Alzheimer’s disease (AD) in which the biology of the disease can be explored during the presymptomatic phase of the illness. The retina is an outgrowth of the central nervous system and therefore provides the opportunity for direct observation of neural tissue and its vasculature during life. Retinal thinning measured *in vivo* has been previously described in persons carrying ADAD mutations through fundoscopy but its pathologic correlates have not been reported.

We describe retinal lesions detected using fundoscopy *in vivo* in a patient homozygous for the A431E mutation in *PSEN1* and its pathological correlates. Retinal lesions seen with fundoscopy during life corresponded to intraretinal and prelaminar optic nerve head amyloid β_42_-protein that were surrounded by perivascular anti-11A50-B10-Aβ_40_ and gliosis. We then performed a cross-sectional, observational study of forty-one Latinos in three cohorts consisting of (1) persons with ADAD causing mutations, (2) at 50% risk for, but testing negative for ADAD mutations, and (3) elderly subjects not at-risk for ADAD.

Clinical exam demonstrated novel, yellow, intraretinal lesions in Cohort 1 in absence of drusen. Fifty-six percent of Cohort 1 subjects had >10 retinal lesions compared to 0% and 25% for Cohorts 2 and 3, respectively (*P* < 0.04). There has been some controversy as to the detectability of Aβ in the retina of persons with AD during life and our findings verify the presence of intraretinal, prelaminar, and perivascular amyloidosis detectable during life in a subset of AD patients.

## Introduction

Alzheimer’s Disease (AD) is characterized by extracellular deposition of Aβ in the brain parenchyma as plaques, and within the walls of small cerebral vessels as cerebral amyloid angiopathy (CAA).^1^ Also present are abnormal intraneuronal cytoskeletal structures containing hyperphosphorylated tau protein as neurofibrillary tangles (NFTs). Histopathological studies of the eyes of patients have demonstrated a reduction in the number of retinal ganglion cells, reduced thickness of the retinal nerve fiber layer and reduction of axonal profiles in the optic nerve of subjects with mild cognitive impairment (MCI) due to AD as well as persons with AD dementia.^2–9^ Thinning of various retinal layers has also been demonstrated *in vivo* in numerous human clinical studies.^7,10–11^ Other *in vivo* studies in humans have shown either impaired arteriolar vessel reactivity, reduced foveal avascular zone size, or decreased blood flow in the superficial macula in late onset Alzheimer’s disease (LOAD).^12–14^ Histopathological analyses of the human retina in LOAD have demonstrated the microscopic presence of tau-related changes^15–18^ as well as beta-amyloid (Aβ) plaques and vascular Aβ deposits.^1,5–7,17–22^ Although histopathological findings and accompanying *in vivo* retinal amyloid imaging reports have demonstrated promising insights,^5–9,15–43^ the clinical identification of AD-specific retinal pathology remains challenging. This difficulty arises from the high prevalence of comorbid retinal conditions such as drusen or hard exudates (e.g. age-related macular degeneration, diabetic retinopathy, etc.) which are indistinguishable from potential AD-related changes during routine clinical examinations in the elderly population.

Unlike the common form of AD (LOAD), around 1% of AD cases are due to fully penetrant genetic mutations that are inherited in an autosomal dominant manner and termed autosomal dominant AD (ADAD).^44^ For subjects that carry an ADAD mutation, development of clinical AD is essentially certain, with the age of symptom onset being relatively young (typically < 60 years of age) and fairly predictable based on the specific mutation present.^45^ Though the clinical and pathological phenotypes of ADAD are generally similar to those of LOAD, there are some differences^46^ including a higher prevalence of CAA in ADAD.^47^ Since ADAD occurs much earlier than LOAD, ADAD subjects serve as a useful model for identifying AD biomarkers by minimizing the influence of confounders that are associated with aging such as age-related macular degeneration, vascular disease and hypertension. As such, characterization of retinal changes in persons with ADAD mutations is a valuable model for understanding the disease course and utility of ocular biomarkers in a “pure” form of AD. Initial studies in ADAD cohorts have found marginally decreased thickness of several retinal layers in presymptomatic *PSEN1* mutation (E280A) carriers^10^ as well as carriers of other mutations^48^ and corresponding attenuation of optic tract projections.^31^ Histological analysis of postmortem retinas from ADAD patients, heterozygous carriers of the *PSEN1* A260V or A431E mutations, revealed elevated Aβ_42_and Aβ-oligomer levels, comparable to those observed in the retinas of LOAD patients.^6^ Abnormally elevated and heterogeneous retinal capillary blood flow patterns have also been demonstrated in presymptomatic carriers of *PSEN1* and *APP* mutations.^49^

Here, we report the first clinical and postmortem histopathological characterization of intraretinal findings in a patient who was homozygous for a pathogenic *PSEN1* (A431E substitution)^50–52^ mutation. We correlate these dilated fundus examination findings with clinical findings on fundus photos, optical coherence tomography (OCT), and OCT angiography (OCTA), and demonstrate that the retinal abnormalities seen *in vivo* correlate spatially with areas of positive intraretinal Aβ staining seen post-mortem. We also report the clinical and ophthalmological findings from a larger cohort of persons with ADAD mutations relative to a cohort of non-mutation carrying controls and a cohort of elderly persons with various levels of cognitive impairment.

## Materials and methods

### Subject Recruitment

This was a cross-sectional study containing three cohorts of Latino subjects recruited in an ongoing NIH funded observational study (R01AG062007) from 2017 to 2022. Cohort 1 consisted of consecutive subjects with genetic mutations known to cause ADAD. Cohort 2 subjects were at-risk members of families in whom ADAD mutations were known to occur but who were themselves not found to be mutation carriers on genetic testing. Cohort 3 subjects were elderly Latino subjects (>50 years old) with various degrees of cognitive impairment. All participants underwent standardized clinical and cognitive evaluations using the Uniform Data Set 3 (UDS3) of the Alzheimer’s Disease Center network^53^ and participants in Cohort 3 underwent amyloid-PET scans using florbetaben to evaluate for the presence of cerebral amyloidosis.

### Ophthalmologic Examination

All subjects underwent a comprehensive ophthalmologic examination including assessment of standardized Early Treatment of Diabetic Retinopathy Severity (ETDRS) visual acuity, intra-ocular pressure, slit-lamp examination and dilated funduscopic examination by a board-certified ophthalmologist and fellowship-trained retina specialist.

### Ophthalmic Imaging

Ophthalmic imaging was performed on both eyes of all subjects after pharmacologic pupil dilation. Cooperation of some symptomatic subjects was limited due to dementia. Color fundus photographs were obtained from up to seven ETDRS fields with a 30-50° true-color fundus camera (TRC 50DX, Topcon, Livermore, CA) and evaluation of the retinal periphery and macula was also performed with pseudocolored wide-field scanning laser fundus camera (Optomap, Optos, Marlborough, MA, USA). Images were reviewed for quality and presence of lesions by trained graders masked to the underlying clinical condition and genotype. Grading was performed for the appearance of the neurosensory retina, retinal vessels, optic nerve, and ocular media as well as any abnormal retinal or choroidal lesions. Fundus photographs which were not of sufficient quality for assessment of lesions were discarded but all images from all recruited subjects were evaluated. Fundus photographs that were partially useful due to vignetting or other media opacity were evaluated for lesions only in the usable portion of the image.

Optical coherence tomography (OCT) images of the central macula (6×6mm) were acquired using a Heidelberg Spectralis (Spectralis, Heidelberg Engineering, Franklin, MA, USA). Retinal thickness and sublayer thickness (nerve fiber layer, ganglion cell layer, inner plexiform layer, inner nuclear layer, outer plexiform layer, outer nuclear layer and outer retinal banding pattern) for each eye were quantified using the commercially available segmentation and measurement software (Spectralis, Heidelberg Engineering, Franklin, MA, USA).

Optical coherence tomography angiograms (OCTA) of the central macula (3×3mm) were obtained using an AngioPlex 5000 (Carl Zeiss Meditec, Dublin, CA, USA). OCTA were automatically segmented by the manufacturer’s commercially available software to construct *en face* superficial retinal layer (SRL) images. The SRL extends from the inner limiting membrane (ILM) through the inner plexiform layer (IPL), which is approximated as 70% of the thickness between the ILM and outer plexiform layer (OPL). This segmentation methodology is commercially available and has been described before.^54^ All images were assessed for quality including signal strength index (provided by manufacturer software), motion artifact, media opacity and foveal centration.

### Quantitative Capillary Perfusion and Density Measures

Quantitation of OCTA images for capillary density and morphology in the SRL was performed as described previously^54–56^ with modifications to exclude the foveal avascular zone (FAZ) and non-capillary vessels such as arterioles and venules. Capillary density was measured as vessel skeleton density (VSD) which represents the total length of capillaries. VSD is calculated by reducing the width of binarized vessels to one pixel and then dividing those skeletonized vessel pixels by the total dimensions of the image excluding areas occupied by non-capillary vessels. Capillary blood flow was measured as capillary flux (cF), which is calculated by averaging the nonbinarized decorrelation intensity values of the pixels occupied by capillaries.^57–58^ This approximates capillary blood flow because the raw decorrelation intensity values for capillaries in an OCTA image are correlated with the number of erythrocytes passing through a capillary segment per unit time.^58^ Capillary density and flux for the autopsied subject were compared against a set of previously published^49^ subjects at risk for ADAD.^19,21,23–28,49^

## Ocular Histopathology

### Processing of eye tissue, retinal flat mounts and cross-sections, and optic nerve

The donor eyes with the optic nerves were collected within 8 hours after time of death and were punctured and fixed in 10% neutral buffered formalin (NBF) and stored at 4°C. The fixed eyes were dissected to create eyecups. The complete neurosensory retinas were isolated, detached from the choroid and sclera, flat mounts prepared, and the vitreous humor thoroughly removed manually, as previously described.^5–6^ The flat mount retina topographical quadrants (superior, temporal, inferior, nasal) were defined by identifying the macula, optic disc, and blood vessels. Flat mount strips (∼2mm wide) were prepared from four predefined geometric subregions: superior-temporal (ST), inferior-temporal (IT), inferior-nasal (IN), and superior-nasal (SN) retina, spanning diagonally from the optic disc to the ora serrata. Fixed retinal strips were processed for cross-sectioning. Each strip measured approximately 2.5cm from the optic disc to the ora serrata, and the central (C), mid-(M), and far (F) peripheral subregions were further defined based on their radial distance from the optic disc. The flat mount temporal quadrant was separated for immunohistochemistry. This tissue preparation technique allowed for extensive and consistent access to retinal quadrants, layers, and pathological subregions.

### Paraffin-embedded retinal cross-sections

Flat mount-derived strips were initially paraffinized using the standard techniques. Next, strips were embedded in paraffin after flip-rotating 90° horizontally. The retinal strips were sectioned (7-10µm thick) and placed on microscope slides treated with 3- aminopropyltriethoxysilane (APES, Sigma A3648). Before immunohistochemistry, the sections were deparaffinized with 100% xylene twice (10 min each), rehydrated with decreasing concentrations of ethanol (100% to 70%), and washed with distilled water followed by PBS.

### Immunohistochemistry

The flat mount temporal quadrant and the deparaffinized retinal cross-sections (tissues) were washed with 1xPBS and treated with target retrieval solution (pH 6.1; S1699, DAKO) at 99°C for 1 hour and washed with 1xPBS. The retinal cross-sections were then treated with formic acid 70% (ACROS) for 10 minutes at room temperature (RT) and washed with PBS before staining for Aβ and pTau burden. For peroxidase-based immunostaining, tissues were washed with wash buffer (Dako S3006) and added 0.2% Triton X-100 (Sigma, T8787) for 1 hour and then treated with 3% H_2_O_2_ for 20 minutes at room temperature and rinsed with wash buffer.

Retinal flat mount temporal quadrant was incubated with mouse anti-human Aβ_42_ monoclonal antibody (12F4 clone; react to the C-terminus of Aβ and specific to Aβ_42_ isoforms; 1:500; Biolegend, Cat.# 805501), for 48 hrs at 4°C. The primary antibody was diluted in the Ab diluent solution with background-reducing components (DAKO S3022). Retinal cross-sections were incubated with 12F4 mAb (1:500) or with rabbit anti-Tau(pSer396) pAb (AS-54977; 1:2000), diluted with background-reducing components, for overnight at 4°C. Tissues were rinsed thrice (10 min each) with wash buffer on a shaker, and incubated for 45 minutes at 37°C with secondary antibodies (anti-mouse Ab, HRP conjugated, DAKO Envision K4001 or anti-rabbit Ab, HRP conjugated, DAKO Envision K4003), and rinsed thrice with wash buffer. For all tissues, 3,3-diaminobenzidine (DAB) substrate was used (DAKO K3468). For retinal cross-sections that were stained with Aβ_42_ mAb, hematoxylin counterstaining was performed. All tissues were mounted with Paramount aqueous mounting medium (Dako, S3025).

For fluorescence-based immunostaining, retinal cross-sections were treated with blocking solution (DAKO X0909) supplemented with 0.1% Triton X-100 (Sigma, T8787), then washed with 1xPBS prior to overnight incubation at 4°C with the following primary antibodies: mouse anti-human Aβ_42_mAb (12F4 clone, 1:200; Biolegend, Cat.# 805501), mouse anti-human Aβ_40_ mAb (11A50-B10 clone; react to the C-terminus of Aβ and specific to Aβ_40_ isoforms; 1:200; Biolegend, Cat.# 805401), rat anti-GFAP mAb (1:500; Invitrogen, Cat. # 13-0300), rabbit anti-collagen IV pAb (1:500; Abcam, Cat. # ab6586); goat anti-IBA1 pAb (1:500; Novusbio, Cat.# NB100-1028). The following day, retinal cross-sections were washed thrice with 1xPBS and incubated with secondary antibodies (1:200; Donkey anti-mouse,-rat,-rabbit or-goat, Cy5, Cy3 and Cy2, Jackson Immunoresearch laboratories, INC.), for 1 hour at room temperature. After rinsing thrice with 1xPBS, sections were mounted with Prolong Gold antifade reagent with DAPI (Thermo Fisher #P36935). Routine controls were processed using identical protocol while omitting the primary antibodies to assess nonspecific labeling.

### Microscopy

Bright field and fluorescence images were acquired using a Carl Zeiss Axio Imager Z1 fluorescence microscope with ZEN 2.6 blue edition software (Carl Zeiss MicroImaging, Inc.) equipped with ApoTome, and AxioCam MRm and AxioCam HRc cameras. Multi-channel image acquisition was used to create images with multiple channels. Tiling mode and post-acquisition stitching were used to capture and analyze large areas. Images were repeatedly captured at the same focal planes with the same exposure time. Images were captured at 20×, 40×, 63×, and 100× objectives for different purposes.

### Group Comparisons

In order to ascertain whether detectable retinal abnormalities associated with ADAD mutation status were present during life in persons with ADAD mutations, fundus photographs from all cohorts were reviewed by a grader masked to the participants’ genetic status and clinical exam findings. The images were graded to determine the presence, number, and location of abnormal retinal lesions within or around the retina, optic nerve head and retinal vessels. These lesion counts were tabulated in relation to disease stage as determined by the Clinical Dementia Rating scale (CDR), presence vs. absence of the pathogenic ADAD mutations, retinal location and age-adjusted for mutation-specific age of dementia diagnosis. As age of disease onset differs more between mutations than within a mutation,^45^ a mutation-specific adjusted-age of diagnosis can be calculated as an estimation of the time remaining from the expected age at which dementia will develop. Comparisons were made between groups using non-parametric Kolmogorov-Smirnov tests and Chi-squared tests where appropriate using R version 4.2.3 (R Foundation for Statistical Computing; Vienna, Austria).

## Data availability

Data from this manuscript are available for review based on written requests for non-commercial use, with appropriate IRB approval and legitimate research goals. Since retinal images are considered inherently identifiable and there are very few subjects with or at risk for ADAD some images may not be sharable to protect subject privacy.

## Results

### Study Population

Of the 25 subjects at risk for ADAD, *PSEN1* and *APP* mutations were found in 15 and three subjects, respectively (Table 1). Among 18 mutation carriers, five were asymptomatic (CDR = 0), three were mildly symptomatic (CDR = 0.5) and 10 had dementia (CDR > 0.5). Among the 18 mutation carriers, two had the F388S substitution in *PSEN1*,^60^ three had the V717I substitution in *AP*P,^61^ and 13 had the A431E *PSEN1* substitution.^51–52^ Cohort 3 consisted of 16 Latinos of whom six had grossly normal cognition, four had mild cognitive impairment, and six had dementia. All six with dementia had positive amyloid-PET scans and were considered to have AD. One person with normal cognition had a positive amyloid-PET scan.

**Table 1.**
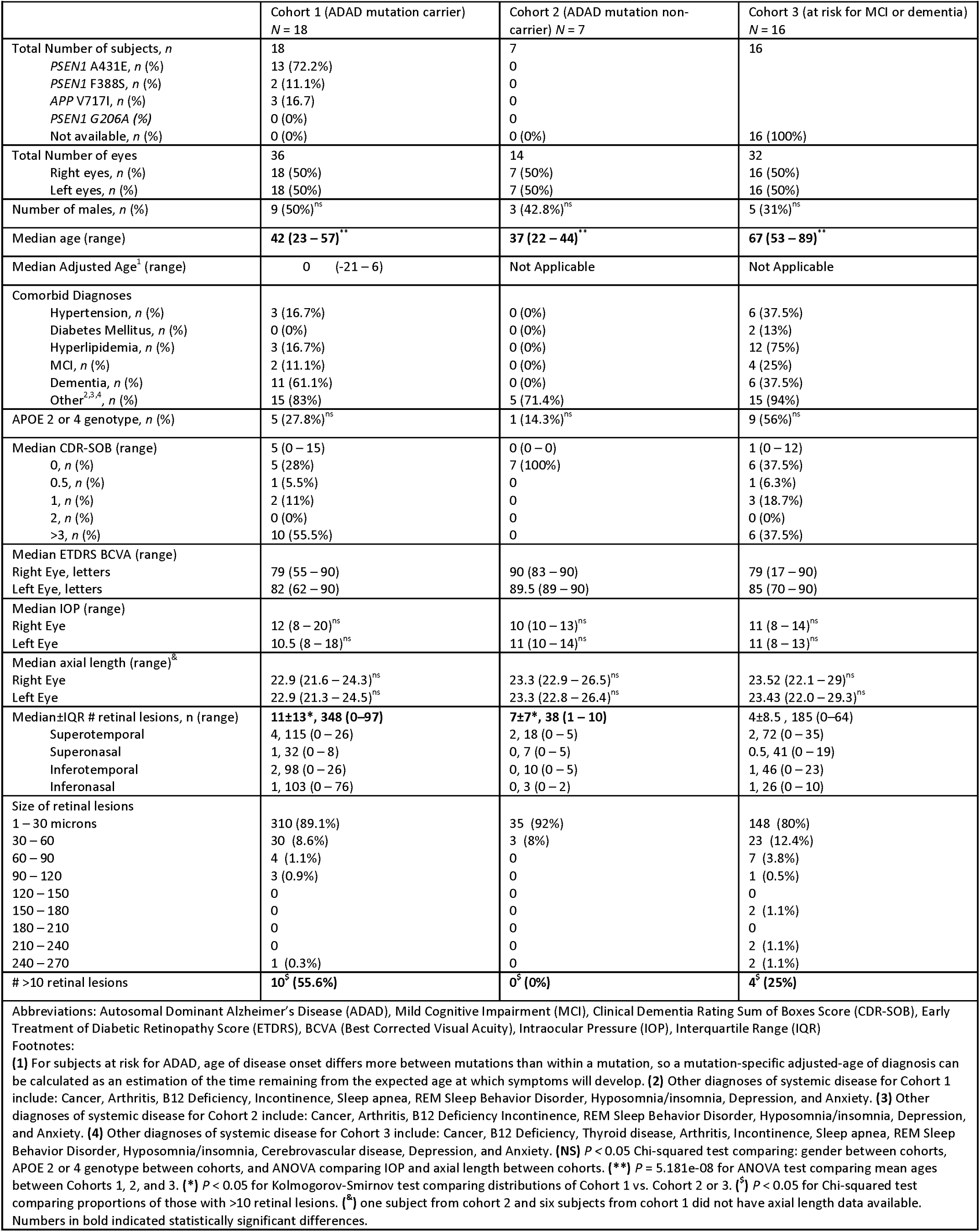
Summary of subject characteristics and ophthalmic findings.

As the *APOE* epsilon 2 and 4 alleles have been found to be associated with CAA,^62^ we assessed these genotypes as well. One out of seven (14%) non-carriers (Cohort 2), and 5/18 (28%) carriers in Cohort 1 had an *APOE* 2 or 4 allele. Of these five carriers, one (20.0%) was an asymptomatic carrier, one (20.0%) was a mildly symptomatic carrier and three (60%) had dementia. Three subjects in Cohort 3 were homozygotes for these alleles and five were of the 3/4 genotype.

### Pre-mortem and post-mortem evaluation of homozygote for A431E substitution in PSEN1

During the course of this study, one subject in Cohort 1 (henceforth called the index subject) passed away from secondary complications of ADAD approximately 20.5 months after his last study examination. Ocular and CNS tissue from this subject was available for histopathologic evaluation as described below. This subject was homozygous for the A431E substitution in *PSEN1.* He began declining in cognition, motor function, and passed away all in his 30s (exact ages have been redacted due to publication guidelines). His overall clinical presentation excluding ocular findings has been previously described.^63^ While he had family history of ADAD, the exact details are redacted due to publication guidelines to protect patient privacy. At the time of ophthalmologic examination he had significant leg spasticity (Ashworth score of 3/4 in both legs), was unable to walk independently, and was hyperflexic throughout with bilateral Babinski signs. He had advanced dementia with a CDR score of 3 (severe dementia) and a sum of boxes score of 18 (maximal disability). He scored 8/30 on the MMSE and the severity of his dementia precluded more formal neuropsychological testing. He had an earlier onset and more aggressive course of the disease than typically seen in association with his mutation. This subject underwent histopathologic evaluation and, while his ophthalmologic findings were consistent with findings in Cohort 1, they were not included in Table 1 due to his inability to perform some clinical ophthalmological testing.

### Neuropathological Evaluation of homozygote for A431E substitution in PSEN1

The post-mortem interval for brain and ocular tissue collection for the index subject was 8 hours. Mild atrophy was evident in the frontal and temporal lobes with moderately severe atrophy appreciable in the hippocampus. Hematoxylin and eosin staining revealed granulovacuolar degeneration, neuronal loss and a few Hirano bodies. The entorhinal cortex also demonstrated neuronal loss and granulovacuolar degeneration. The neocortices (frontal, temporal, parietal) all demonstrated neuronal loss. Anti-β amyloid immunostain 4G8 revealed frequent, diffuse plaques in the hippocampus, frontal, temporal, parietal, occipital lobes, basal ganglia, pons and midbrain. A moderate amount of plaques were also identified in the cerebellum. Grade I amyloid angiopathy was identified in the hippocampus CA1, entorhinal cortex and in the neocortices (frontal, temporal, parietal and occipital lobes). Gallyas silver stain revealed frequent neuritic plaques, neurofibrillary tangles and neuropil threads in the CA1 region of the hippocampus, entorhinal cortex, frontal, temporal, parietal and occipital cortices. The midbrain showed no Lewy bodies, neuronal loss or gliosis though it did demonstrate pigmentary incontinence. With anti-alpha-synuclein immunostain, there were also no Lewy bodies present in the substantia nigra, locus coeruleus or medulla. The ADNC grading was A3, B3, C3, representing a high probability of Alzheimer’s Disease. The Braak score was VI.

### Ophthalmological Evaluation of the homozygote for A431E substitution in PSEN1

The index patient was unable to cooperate with visual acuity testing due to his advanced dementia. His smooth pursuit eye movements had saccadic intrusions; the full assessment of extraocular motility was limited as he was unable to reliably follow commands. The intraocular pressures were within normal limits for both eyes. The slit-lamp examination of the anterior segment was unremarkable. Dilated funduscopic examination demonstrated multiple, small to very fine, yellow deposits that appeared to be most prominent in the neurosensory retina of both maculae and range in size from 20-100 microns as noted in other subjects in Cohort 1 (Fig. 1). Color fundus photographs from the index subject demonstrated some of the retinal lesions that had been observed clinically although many were not resolved or remained slightly out of focus due to motion artifacts and the subject’s limited ability to position (Fig. 2). Color images of the optic disc demonstrated moderate perivascular whitening.

**Figure 1.**
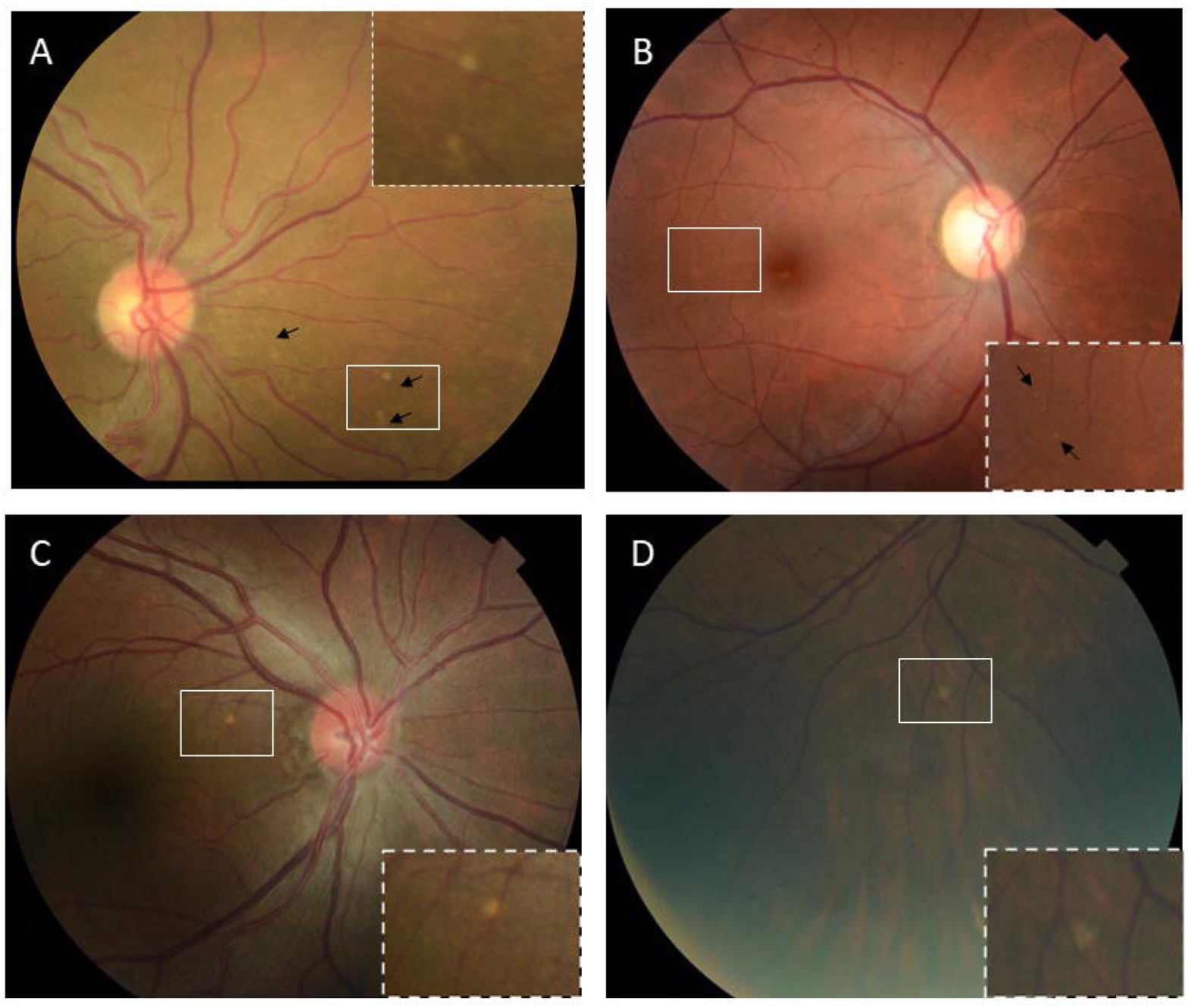
True color fundus photographs from 4 different representative subjects at risk for Autosomal Dominant Alzheimer’s Disease (ADAD, Cohort 1). (A) Subject 1 is a 55-59 year old female with an APP V717I mutation, APOE 3/3 genotype, and adjusted-age of 2. There was no medical history of diabetes mellitus, hypertension or hypercholesterolemia. CDR-SOB score was 10. There were 97 lesions detected on all available fundus images for this subject. In addition to the area of the white box, there were numerous fine lesions immediately nasal to the disc which may be difficult to resolve on the fundus photograph. (B) Subject 2, a 40-44 year old female with PS1 A431E mutation, APOE 2/3 genotype, and adjusted-age of 2. CDR-SOB score was 13. There were 66 lesions detected on all available fundus images for this subject. (C) Subject 3, a 45-49 year old male with PS1 A431E mutation, APOE 3/3 genotype, and adjusted-age of 0. CDR-SOB score was 7. There were 7 lesions detected on all available fundus images for this subject. (D) Subject 4, a 55-59 year old female with no family history of dementia or risk for ADAD. Genotyping for this subject was not available. CDR-SOB score was 0. There were 4 lesions detected on all available fundus images for this subject. Exemplary lesions for all cases are illustrated in white box, inset and/or black arrows. True color images were acquired on a Topcon TRC 50DX.

**Figure 2.**
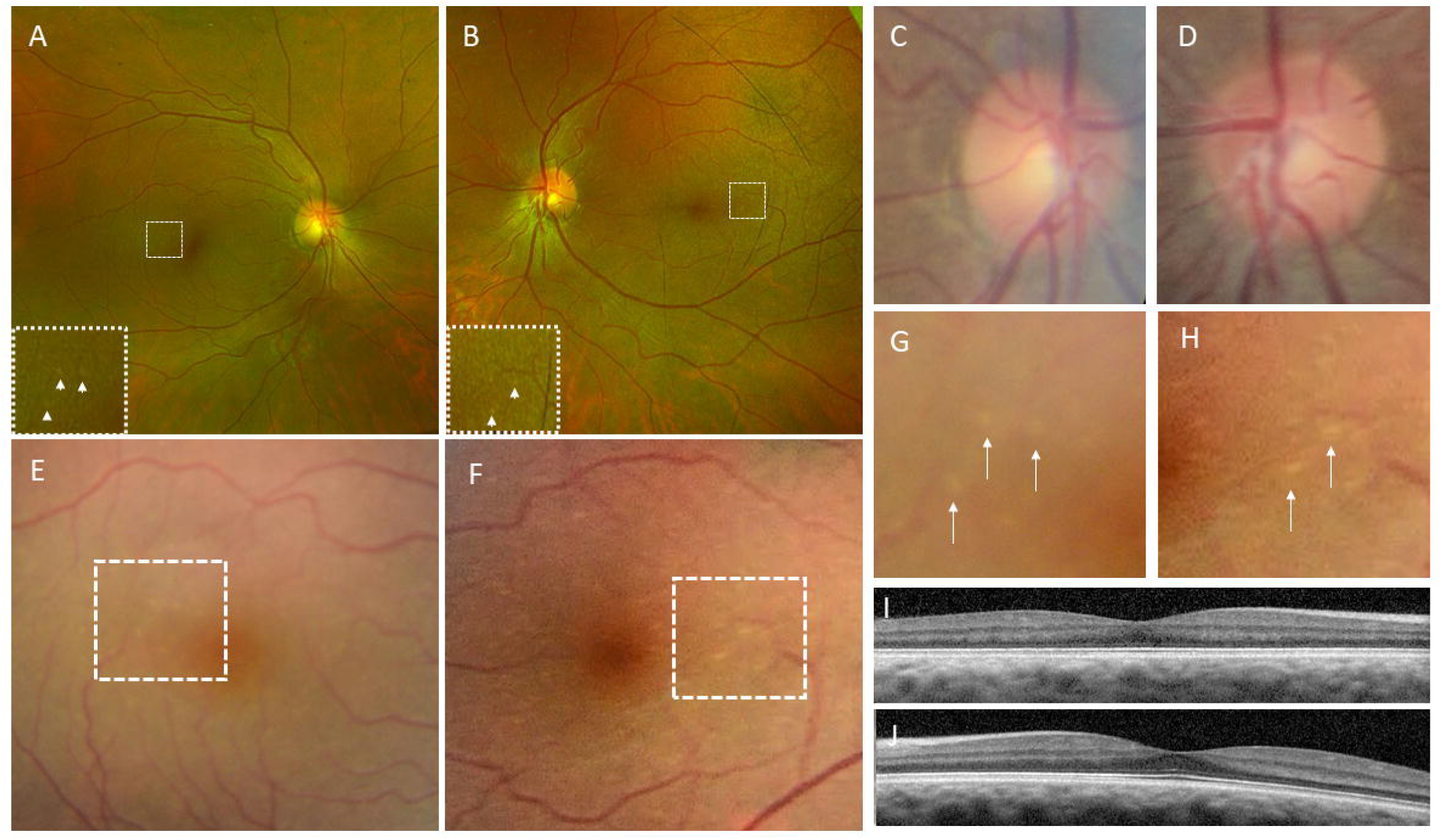
Fundus photographs of an index 35-39 year old PSEN1 homozygote who passed away during the course of the study from complications associated with ADAD. Pseudocolor widefield fundus images of the posterior pole in the. **(A)** right and **(B)** left eyes demonstrate subtle fine yellow lesions in the macula that were initially noted on clinical examination. Higher magnification images are provided in inset. White arrowheads indicate locations of some of the more prominent clinically visible lesions. An incidental choroidal nevus is noted in the superior macula. **(C)** True color fundus photograph of the right optic disc and **(D)** left optic disc demonstrate very mild perivascular whitening along the nasal artery. **(E)** Magnified true color fundus photograph of right macula taken on a true color fundus camera more clearly demonstrates fine refractile, yellow lesions in temporal macula. **(F)** Similar findings in magnified true color images of left macula. **(G)** Magnified region of white box from panel (E). **(H)** Magnified images from white dotted box from panel (F). Foveal OCT B-scans of the **(I)** right eye and **(J)** left eyes show unremarkable foveal depression and retinal contour. There were no drusen on any macular OCT scan noted in this subject. White arrows and arrowheads in all panels indicate location of exemplary and clinically notable lesions. Many more lesions were visible on clinical examination with 2.2 diopter and 66 diopter lenses than are apparent on fundus photographs due to subtle blurring from patient motion. Pseudocolored images were taken on an Optos 200Tx. True color images were acquired on a Topcon TRC 50DX. All images were acquired 20.5 months before death. Images have not been contrast altered or visually enhanced.

OCT scans of macular thickness in the index subject demonstrated normal foveal contour and no clear intraretinal or subretinal lesions in any section (Fig. 2). Specifically, no retinal drusen were noted on any OCT section in any subject in Cohort 1. We compared the total retinal thickness and sublayer thickness of the index subject to corresponding published mean retinal measurements from subjects carrying other ADAD causing mutations and controls.^10,48–49^ Total retinal thickness as well as all sublayer thicknesses (retinal nerve fiber layer, ganglion cell layer, inner plexiform layer and outer nuclear layer) for the index subject were well below published values from subjects with similar mutations and controls consistent with the advanced stage of the index subject’s disease (Table 2). OCTA of each eye did not demonstrate qualitatively obvious focal areas of capillary loss (Supplemental Fig. 1A,B). However, capillary perfusion was both qualitatively and quantitatively lower (Supplemental Fig. 1D). Both capillary blood flow and capillary density metrics for the index subject were more than one standard deviation below the mean of a group of 33 eyes from 17 normal subjects (Supplemental Fig. 1E,F). The average age of this group of normal subjects used for comparison was 37.8 ± 10.3.

**Table 2.** Retinal Thickness for Index Subject with PSEN1 Mutation Compared to Other Similar Reported Mutations and Controls From Literature.

|  |  |
| --- | --- |
| Total Retinal Thickness (microns)<br>Index Subject OD Average<br>Index Subject OS Average<br>PSEN1E280A Non-carriers<sup>30</sup><br>PSEN1E280A Carriers<sup>30</sup><br>Adhikari et al.,<sup>22</sup> | 324<br>331<br>344±16<br>330±16<br>337±13 |
| RNFL (microns)<br>Index Subject OD Average<br>Index Subject OS Average<br>PSEN1E280A Non-carriers<sup>30</sup><br>PSEN1E280A Carriers<sup>30</sup><br>Adhikari et al.,<sup>22</sup> | 21<br>25<br>23±4<br>23±4<br>27±2 |
| GCL (microns)<br>Index Subject OD Average<br>Index Subject OS Average<br>PSEN1E280A Non-carriers<sup>30</sup><br>PSEN1E280A Carriers<sup>30</sup><br>Adhikari et al.,<sup>22</sup> | 42<br>44<br>52±6<br>51±8<br>46±5 |
| IPL (microns)<br>Index Subject OD Average<br>Index Subject OS Average<br>PSEN1E280A Non-carriers<sup>30</sup><br>PSEN1E280A Carriers<sup>30</sup><br>Adhikari et al.,<sup>22</sup> | 37<br>37<br>41±3<br>42±4<br>40±4 |
| ONL (microns)<br>Index Subject OD Average<br>Index Subject OS Average<br>PSEN1E280A Non-carriers<sup>30</sup><br>PSEN1E280A Carriers<sup>30</sup><br>Adhikari et al.,<sup>22</sup> | 66<br>61<br>75±12<br>67±9<br>87±8 |
| Retinal thickness values for index subject and comparator values. Total retinal thickness and sublayer thickness values for index subject's left and right eyes were obtained from the average of the central subfield and surrounding subfields with the 3mm ETDRS grid using commercially available segmentation software (Heidelberg Spectralis). Comparator average values from recent publications are listed for both eyes of subjects carrying known ADAD causing mutations or controls. RNFL = Retinal Nerve Fiber Layer. GCL = Ganglion Cell Layer |  |

### Elevated Aβ_42_, pTau, vascular Aβ_40_, and gliosis in ADAD retina

Both donor eyes were harvested from the index subject approximately 20.5 months after his last clinical evaluation. Immunohistochemistry of retinal flat mounts from the left eye using anti-Aβ_42_ monoclonal antibody and previously described methods^20^ demonstrated diffusely and densely positive staining in regions of the retina that had been noted before to have clinically visible retinal lesions. Using retinal vessels as landmarks, focal areas of increased immunostaining were noted and corresponded to clinically visible retinal lesions on fundus images and examination (Fig. 3).

**Figure 3.**
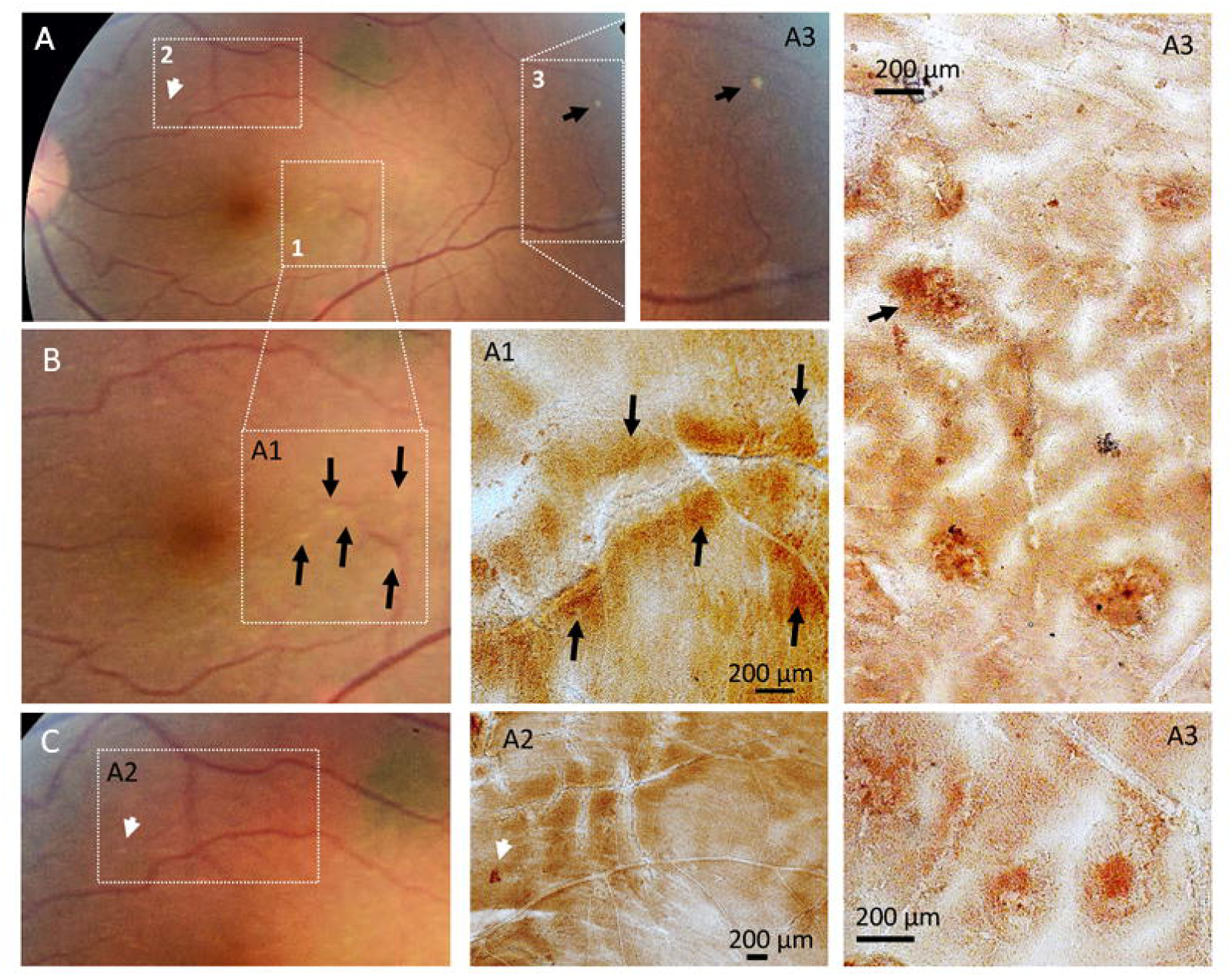
Histopathology of. **A**β**_42_ deposition in flat mount of retinal regions and corresponding fundus images from an index subject with PSEN1 mutation. This is the same subject as illustrated in** Fig. 2**. (A)** True color fundus photograph of subject’s left eye. An incidental choroidal nevus is noted in the superior macula. Regions labelled 1-3 in white dotted boxes represent areas with exemplary clinically visible lesions although numerous other regions with fine lesions were also notable on clinical examination. The most easily evident of these lesions is in area 3. **(B,C)** Higher magnification of regions in panel A. White or black arrows highlight the location of lesions. Immunohistochemistry for Aβ_42_, labelled by using the 12F4 monoclonal antibody, on retinal flat mounts from the numbered regions in the white dotted regions is provided in subsequent panels A1-A3. Additional image from the A3 flat mount region included several large Aβ plaques. **(A1)** Region temporal to the fovea was noted to have numerous fine refractile, yellow lesions near a retinal vessel (red) coming in from the temporal region. Translucent sharp lines represent retinal blood vessels in flat mounts and provide orientation and landmarks.

Retinal flat mounts were cut and cross-sectioned from the superior-temporal and inferior temporal regions were prepared as described in the methods section to evaluate the presence of AD-related pathology, including Aβ and pTau reactivity within the retinal layers (Fig. 4). Retinal cross-sections throughout the superotemporal region of the left eye demonstrated diffuse and dense anti-Tau (pSer396) immunoreactivity that was most prominent in the inner plexiform layer and less prominent in the outer nuclear layer (Fig. 4A-B). Numerous cell bodies in the inner nuclear layer and surrounding inner blood vessels were exhibiting intense pS396-tau inclusions as well (Fig. 4C-G). There was diffuse reactivity throughout the retinal cross-section for anti-Aβ_42_and this was most prominent in the nerve fiber layer and especially in regions surrounding blood vessels (Fig. 4H-J). Immunofluorescence for Aβ_42_ (12F4), along with vascular collagen IV and GFAP-positive astroglia showed the presence of Aβ_42_ deposits within the retinal nerve fiber layer and within the walls of larger retinal vessels confirming the presence of retinal vascular amyloidosis (Fig. 5A-B). Moreover, dense vascular Aβ_40_ (11A50-B10) signals within collagen IV-positive vessel walls and multiple activated IBA-1-positive microglia were observed in the retina of this ADAD patient (Fig. 5C).

**Figure 4.**
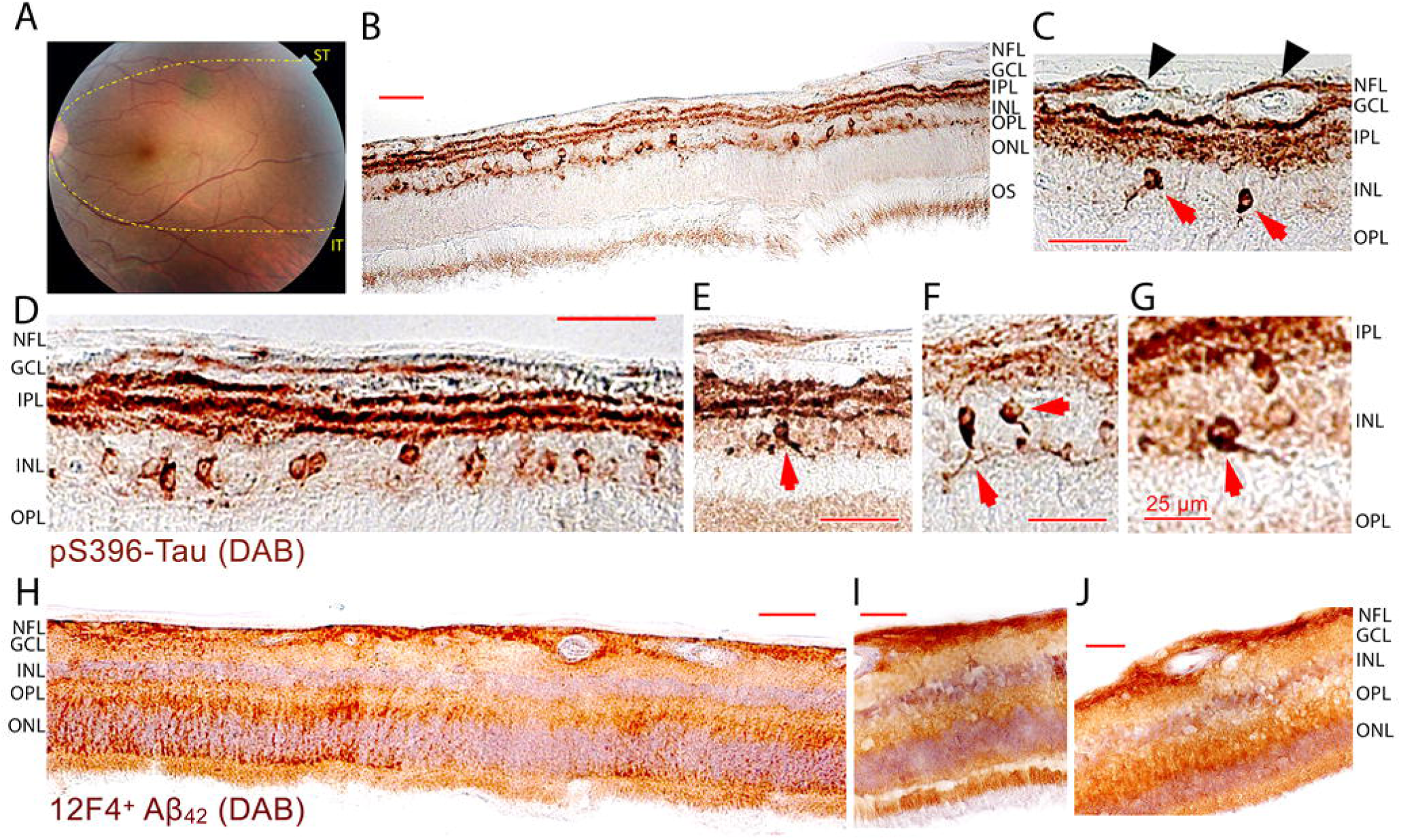
Retinal A**β**_42_ and hyperphosphorylated tau (pSer396) immunoreactivity in cross-sections from a 35-39 year old ADAD patient homozygous for A431E mutation in PSEN1 gene. Immunohistochemical analysis of A**β**_42_ (12F4) and Tau(pSer396) immunoreactivity in retinal cross-sections from 35-39 year old index PSEN1 homozygote. Retinal cross-sections were prepared as described in the methods section. **(A)** Color fundus photograph of the left eye (OS). Yellow lines show the superior-temporal (ST) and inferior-temporal (IT) strips from which retinal cross-sections were prepared for immunostaining. **(B-G)** Anti-Tau(pSer396) immunoreactivity was visualized by peroxidase-based DAB labeling and noted most prominently in the inner plexiform layer and in numerous cell bodies located in the inner nuclear layer. **(C)** Dense retinal pTau signals surrounding inner blood vessels (black arrowheads) within the NF/GC layers. **(D)** Intracellular pTau accumulation within INL cells and along the IPL synapses and ganglion cell axons. **(E-G)** Higher magnification images showing NFT-like structures of dense pTau inclusions (red arrows) within INL cells. **(H-J)** Anti-Aβ_42_ immunostaining using the 12F4 mAb and visualized by peroxidase-based DAB labeling showed the presence of diffuse Aβ_42_deposits across all retinal layers and most densely in the retinal nerve fiber layer. More intense staining was noted around and within retinal vessel walls. Occasionally, intense Aβ_42_ staining is further found in the outer retina, in photoreceptors, above the outer limiting membrane, and in the IPL. Scale bars = 50 µm, unless otherwise specified.

**Figure 5.**
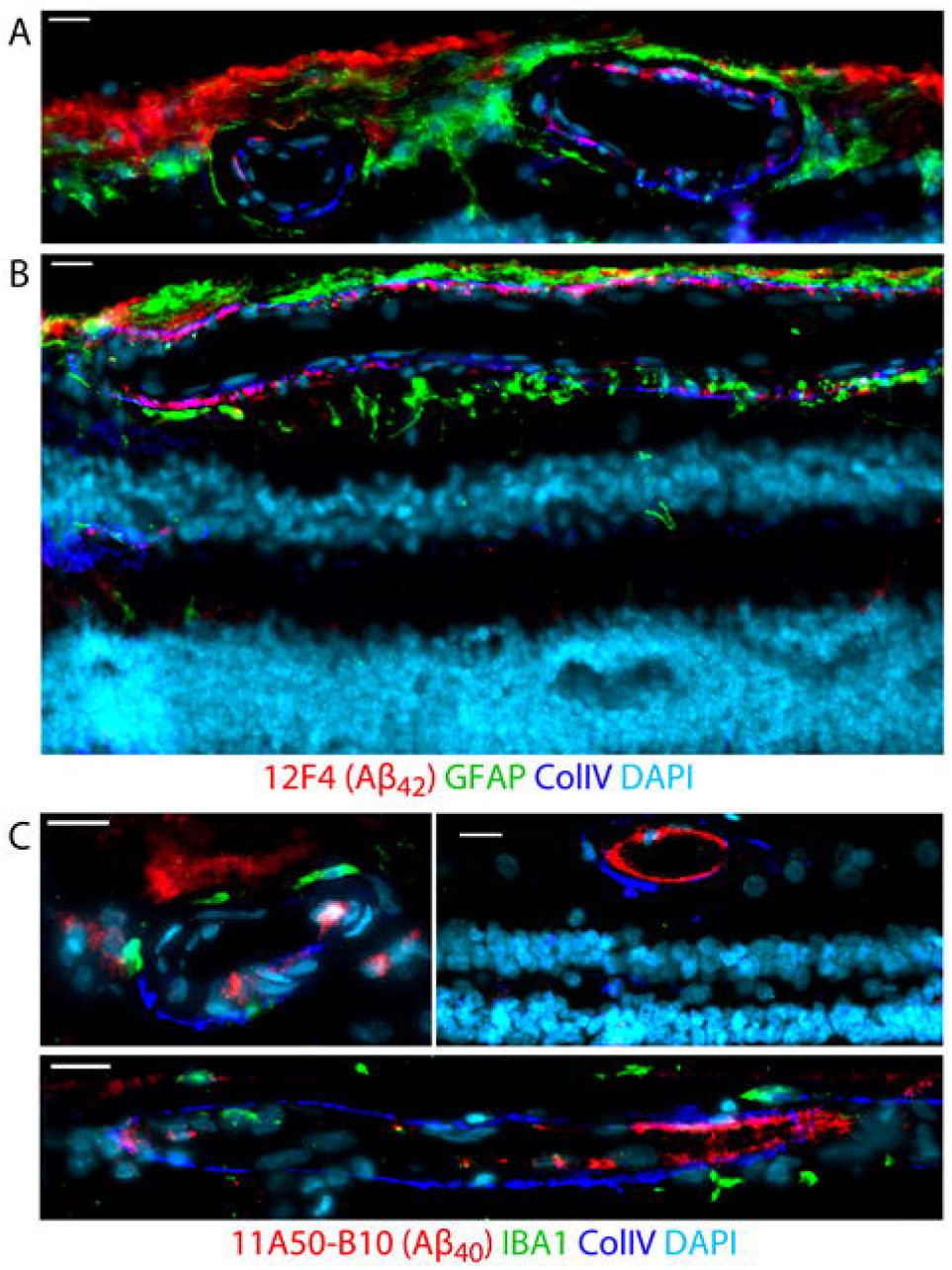
Retinal vascular amyloidosis and gliosis in 35-39 year old index PSEN1 homozygote. (A,B) Representative images of immunofluorescence staining for Aβ_42_ (12F4, red), GFAP astrocytes (green), vascular collagen IV (CollV, blue) and nuclei (DAPI, cyan) in retinal cross-sections from the index ADAD patient. Intense Aβ_42_ accumulation appears around and within inner blood vessels and the nerve fiber layer (NFL). **(C)** Representative images of immunofluorescence staining for vascular Aβ_40_ (11A50-B10, red), IBA1 microglia (green), vascular collagen IV (CollV, blue), and nuclei (DAPI, cyan) in retinal cross-sections. Intense Aβ_40_deposition is detected within inner blood vessel walls. Scale bars = 20 µm.

### Histopathology of optic nerve from the index homozygote for A431E substitution in PSEN1

Aβ_42_ was found in the prelaminar (anterior) optic nerve head (Fig. 6). In contradistinction, there was a paucity of Aβ_42_ within the lamina cribosa and almost no Aβ_42_ in the post-laminar optic nerve head where the axons are myelinated. (Fig. 6A-C) The anteriorly located Aβ_42_ appeared in aggregates and sometimes in the vicinity of a few identifiable plaques (Fig. 6D-E). There was also a dense accumulation of Aβ_42_ around the blood vessels of the optic nerve head (Fig. 6C) which corresponded to perivascular whitening on fundus photographs (Fig. 2D). In the retina adjacent to the optic disc, both nasally and temporally, there was very little Aβ_42_.

**Figure 6.**
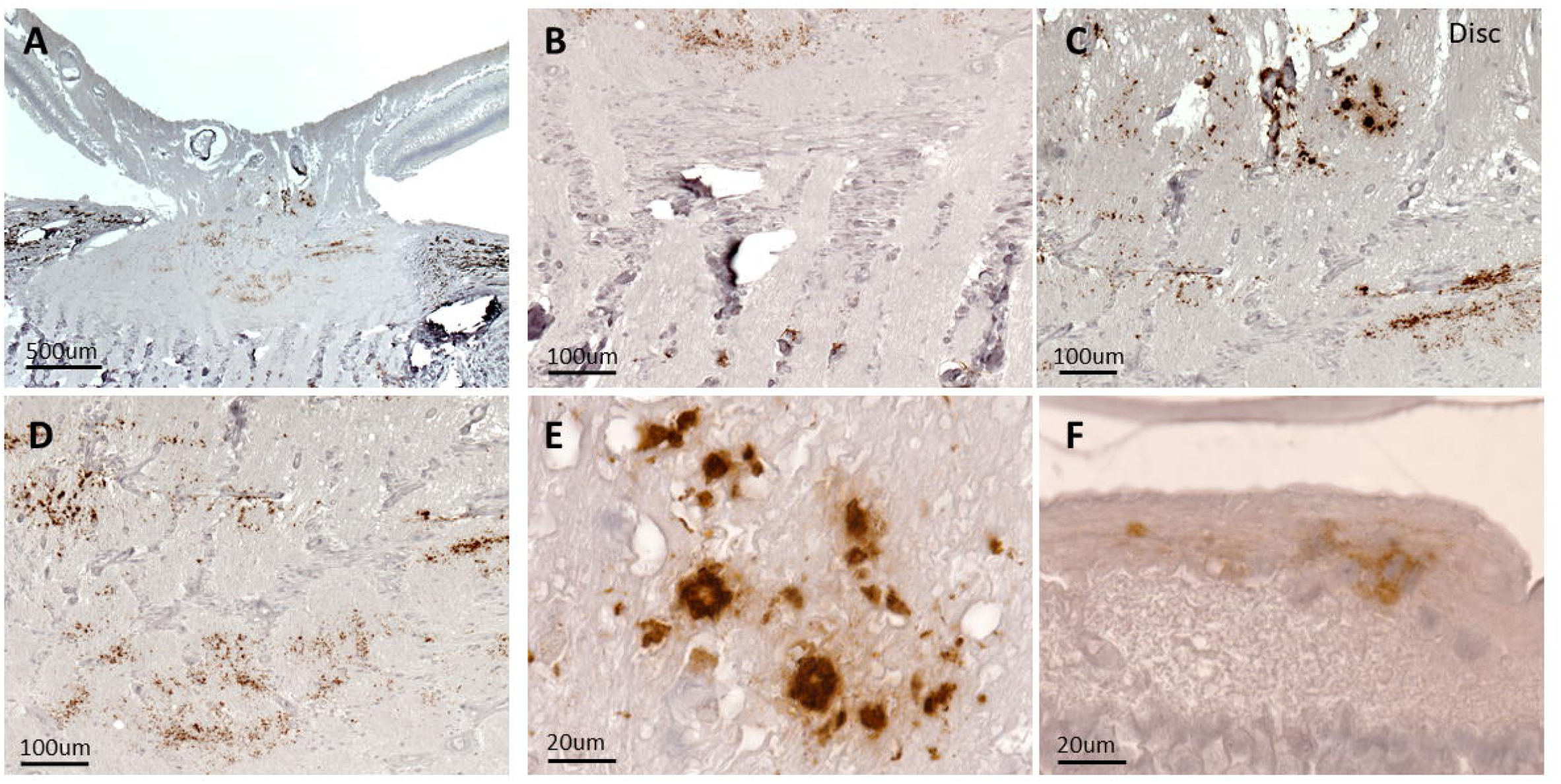
Histopathology of Aβ_42_ deposition in cross-sections of the optic nerve head from index subject with PSEN1 mutation. **(A)** Low magnification image of the optic nerve head shows positive Aβ_42_ staining in the prelaminar space and near a few larger vessels. Higher magnification images demonstrating **(B)** essentially no staining at or posterior to the lamina cribrosa, **(C-E)** numerous areas of Aβ_42_ staining in the prelaminar region and **(F)** modest Aβ_42_staining in the peripapillary retina.

### Retinal lesions on dilated fundus examination across cohorts

Fundus examinations and imaging were available for all 82 eyes from 41 subjects in all cohorts. Comorbid health conditions among the relatively young subjects in Cohort 1 (subjects with mutation predisposing to ADAD) and Cohort 2 (family members with no mutation predisposing to ADAD) were less prevalent compared to the elderly subjects in Cohort 3. Table 1 summarizes the subject demographics. Dilated fundus examination of Cohort 1 subjects demonstrated yellow, mostly circular, retinal lesions of varying sizes that were atypical for the relatively young age and low burden of comorbid disease in this cohort (Fig. 1). Among all subjects in Cohort 1, there were 348 lesions with a median (±IQR) of 11±13 lesions per subject. Among all non-mutation carrying relatives of ADAD subjects (i.e. Cohort 2 subjects), there were 38 lesions with a median (±IQR) of 7±7 lesions per subject. Among all elderly subjects in Cohort 3, there were 185 lesions with a median (±IQR) of 4±8.5 lesions per subject. The number of retinal lesions in Cohort 1 was statistically greater than in Cohort 2 (*D* = 0.56, *P* = 0.04). When comparing Cohort 1 to Cohort 3 the number of total retinal lesions was approaching statistical significance (*D* = 0.40, *P* = 0.08). Using the median number of lesions in Cohort 1 (10 lesions) as a threshold for comparison of the three cohorts, there were no subjects in Cohort 2 that had more than 10 lesions and only 4/16 (25%) subjects in Cohort 3 with more than 10 lesions. In contrast, 10/18 carriers (56%) in Cohort 1 had more than 10 lesions (Chi-squared = 7.8939, *P* = 0.02), supporting the observation that these findings are related to the presence of ADAD mutations.

Among subjects with ADAD causing mutations (Cohort 1), there were 115 (33%), 98 (28%), 103 (30%), and 32 (9%) lesions in the superotemporal, inferotemporal, inferonasal and superonasal quadrants, respectively. Among the non-ADAD mutation carriers in Cohort 2, there were 18 (47.4%), 10 (26.3%), 3 (7.9%), and 7 (18.4%) lesions in similar regions, respectively. Among elderly subjects (Cohort 3), there were 72 (38.9%), 46 (24.8%), 26 (14.1%), and 41 (22.2%) lesions in similar regions, respectively. Seven subjects (39%) in Cohort 1 and four subjects (22%) in Cohort 3 had lesions 2 disc diameters (DD) or closer from the optic nerve head (ONH), while all lesions in Cohort 2 were 3 DD or further from the ONH (Chi-squared = 3.9274, *P* = 0.1403). In terms of size, 89% and 80% of lesions were between ∼1-30 microns in ADAD and controls (Cohort 3), respectively (*D* = 0.385, *P* = 0.06). In a few cases from Cohort 1 and Cohort 3, lesions were irregularly shaped and were greater than 400 microns. There was no direct relationship with the number of retinal lesions and either participants’ adjusted-age nor with disease status as indexed by the CDR sum of boxes.

## Discussion

We describe the clinical examination and diagnostic imaging findings, consistent with intraretinal amyloid lesions, in a cohort of relatively healthy young subjects with mutations predisposing to ADAD. Many of these lesions were perivascular in distribution. Unlike older subjects, these subjects were all below the age of 60 and had fewer comorbid conditions that would predispose to retinal lesions such as drusen, hard exudates or other retinal pigment epithelial pathology. We show the frequency of retinal lesions is higher in subjects with ADAD predisposing mutations when compared to controls at-risk but not carrying such mutations. A similar but insignificant trend is noted when comparing with elderly subjects with cognitive impairment due to other causes. To support the association of intraretinal lesions observed on clinical examination with underlying ADAD pathology, we also show histopathology from the retina and optic nerve head of an index patient from this cohort who was homozygous for the A431E mutation in *PSEN1*. To our knowledge, this is the first paper to report the dilated fundus examination findings, including fundus photos, coregistered with histopathology of intraretinal and pre-laminar disc amyloid deposition. Notably, no subject with an ADAD mutation had drusen on macular OCT scans, including the index subject who had macular lesions. Therefore, we do not believe these lesions are retinal drusen. Previous studies have reported a sluggish pupil flash response among pre-symptomatic carriers of an E693Q amyloid precursor protein (*APP*) mutation.^64^ This is consistent with the findings that melanopsin retinal ganglion cells (mRGCs), which subserve the afferent limb of the pupillary response, are early targets of Aβ pathology in AD.^7^ *APP* mutations, like *PSEN1* mutations, are ADAD causing mutations with the E693Q *APP* mutation causing disproportionate CAA.

The lesions we report on dilated fundus examination and in fundus photos in the index subject were associated with areas found on histopathological analysis to have Aβ deposition within the retina (Fig. 4) and optic disc (Fig. 6). Beyond detection of Aβ plaques in flat mounts, we observed the accumulation of Aβ_42_ and vascular Aβ_40_ deposits, pS396-tau isoforms, and IBA1^+^ micro-and GFAP^+^ macro-gliosis in retinal cross sections from this ADAD patient (Fig. 4-Fig. 6). These retinal pathological changes are consistent with those previously described in LOAD patients.^5–9,15–23,27–43^ Notably, the index patient was already quite symptomatic at the time of his eye examination. As pathological abnormalities associated with AD may occur decades before onset of dementia, an important goal of research into AD pathology in the retina is to develop early biomarkers for AD before the more obvious clinical onset of symptoms. Because the index patient was already symptomatic, it is unclear at what point the retinal lesions would have clinically manifested in this patient. It is also possible that these lesions change, evolve or even resolve with time. Regardless, in this study we provide evidence that the AD hallmark pathology is present in the retina and is amenable to routine color fundus imaging. More specialized forms of retinal imaging may have additional benefits in detecting these lesions.^5,20,65^

Masked assessment of retinal images from 25 persons known to harbor ADAD mutations confirmed that the retinal lesions are ∼1.6 times more abundant among carriers than in a cohort of subjects at risk for inheriting ADAD mutations (family members without mutations), and ∼2.8 times more abundant than in an unrelated cohort of subjects with cognitive impairment and a median age more than 20 years older. Though sometimes present in early disease (e.g. one carrier of the A431E *PSEN1* mutation with a CDR score of 0.5 had 46 lesions), they were not ubiquitous, even in persons with more advanced dementia. The participant with the most lesions (97, affected by the V717I *APP* mutation) had a CDR score of 2.0 and the single patient with a CDR score of 3 (also with the A431E *PSEN1* mutation) had only 11 lesions identified. The longevity of these lesions is also not known and it is possible that they turn over with time. There is substantial heterogeneity in the clinical and neuropathological features between ADAD mutations,^46^ with some having atypical amyloid plaque composition and morphology^60,66^ or having variable predisposition to CAA,^47^ so the relationship between cerebral and ophthalmological Aβ deposition may vary with the mutation.

Many of the retinal lesions we noted appeared to be perivascular or near blood vessels in distribution so it is possible that their appearance in the retina is subject to the same factors determining the presence of CAA. Indeed, both the A431E and F388S mutations in *PSEN1* are associated with a high prevalence of CAA (and atypical “cotton wool” amyloid plaques)^47,60^ and intraretinal lesions may be more likely to occur in these cases. Measures of retinal capillary perfusion in the index subject were consistent with previous descriptions from our group and others.^14,49,67^ Specifically, retinal capillary perfusion was decreased as measured by OCTA compared to a set of previously published subjects at risk for ADAD. Previous studies have also reported that retinal Aβ plaques can be associated with blood vessels^5^ and other studies have demonstrated retinal vascular changes in AD.^12–14^ We have previously reported *increased* retinal capillary perfusion in presymptomatic ADAD mutation carriers and decreased perfusion in late-stage disease.^49^ These data are consistent with there being an initial phase of hyperemia associated with early inflammation or compensatory blood flow that declines as the disease advances.^68–69^ The clinical appearance of retinal lesions may reflect an underlying imbalance in amyloid deposition and clearance related to retinal perfusion changes. Our findings provide more evidence that retinal vascular measurements from OCTA should be evaluated as a potential biomarker for AD.

The study of persons with ADAD allows us to more accurately define changes specific to the AD process and are less subject to influence by the aging process itself or its co-morbidities such as retinal drusen (associated with age related macular degeneration), hard exudates (associated with retinal vascular diseases such as diabetes retinopathy or hypertension) or other retinal lesions. As such, the generalizability of these findings to the characterization of LOAD is unknown as the co-morbidities of aging are associated with confounding non-specific changes in the retina. Methods seeking to label retinal lesions as amyloid in humans *in vivo* are being developed^5,19,23–28^ and, if successful, could change the way AD is currently diagnosed and managed.

An important consideration in this study is the disproportionate number of carriers of the *APOE* epsilon 2 or 4 alleles among ADAD carriers with dementia. As these genetic variants are known to increase the risk of CAA, and therefore possibly of amyloid detectable in the retina, this could have confounded our results. Though the mean number of lesions among carriers of the *APOE* epsilon 2 or 4 alleles, regardless of ADAD mutation status, was numerically higher (15.73 vs. 12.88, *P* = 0.68) this was not statistically significant nor were there significant differences in the number of carriers of *APOE* epsilon 2 or 4 allele carriers that had more than 10 lesions relative to non-carriers of these variants (47% vs. 35%, *P* = 0.67. Further study with larger numbers of participants will be necessary to determine if there are independent or combined contributions of ADAD mutation and *APOE* genotype to these retinal lesions or CAA. Defining the nature and prevalence of these retinal lesions across the AD spectrum will require larger studies employing a wide range of participants with diverse subtypes of the disease.

In conclusion, we show definitive evidence that deposits of Aβ in the retina and optic nerve head can be detected *in vivo* with fundoscopy or clinically available fundus imaging in a homozygote for the A431E mutation in *PSEN1*. Our evidence in a wider population of living persons with ADAD mutations suggests this is a significant feature of this disease and may be an early biomarker of disease burden. The degree and manner to which these findings can be applied to the larger population at-risk for AD remains to be determined and will require further study.

## Acknowledgements

The authors would like to acknowledge Drs. Carol Miller and Debra Hawes for the neuropathological evaluation of the subject. Dr. Amir Kashani performed or supervised the acquisition and analysis of all ophthalmologic data. Dr. John Ringman performed or supervised the acquisition and analysis of all clinical neurological data. Dr. Maya Koronyo performed or supervised the postmortem analysis of the retina. Dr. Alfredo Sadun performed or supervised the postmortem analysis of the optic disc. Dr. Xuejuan Jiang supervised the statistical analyses. The authors would also like to thank Ms. Anna Tang for help with preparation and submission of the manuscript.

## Supplementary material

Supplementary material is available online.

---

**Supplementary Figure 1 Full thickness, pseudocolored, *en face* and foveal centered OCTA scan (AngioPlex, Carl Zeiss Meditec, Inc) of a 35-39 year old PSEN1 homozygote.** 3×3mm image of retinal capillary perfusion in the right **(A)** and left eye **(B).** Quantitation of capillary blood flow **(E)** and vessel skeleton density **(F)** as measured by OCTA of the index patient in red compared to normal subjects in grey with black bars corresponding to one standard deviation above and below the mean for a set of 33 eyes from 17 normal subjects (Singer et al., ADAD 2021). Note that for both measurements, the patient was more than one standard deviation below the mean.

