## Supplementary figures and images for "Retinal and Optic Nerve Lesions Correspond to Amyloid in Autosomal Dominant Alzheimer’s Disease"

### Supplementary Fig

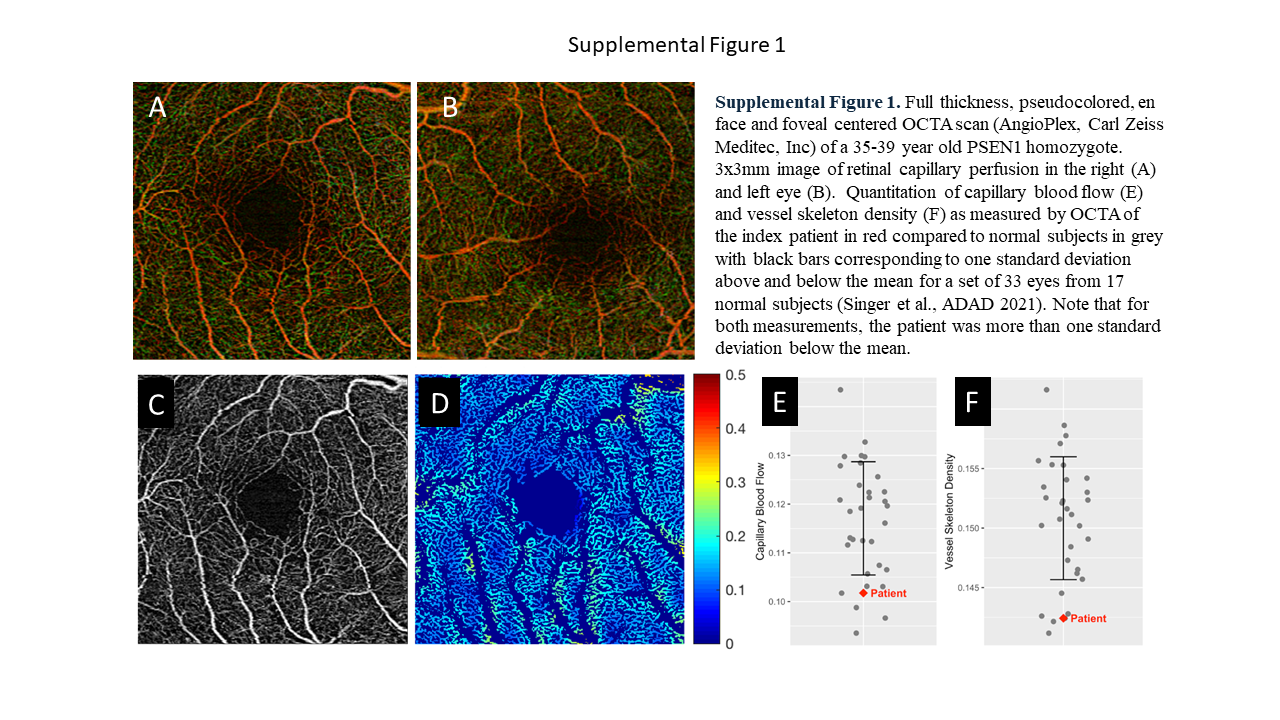
